# Maternal blood lipidomics analyses link critical metabolic pathways associated with severe preeclampsia

**DOI:** 10.1101/2020.07.05.20145292

**Authors:** Yu Liu, Bing He, Mano R Maurya, Paula Benny, Cameron Lassiter, Hui Li, Shankar Subraminiam, Lana X. Garmire

## Abstract

Preeclampsia is a pregnancy specific syndrome characterized by hypertension and proteinuria after 20 weeks of gestation. To reveal the relationship between lipids and preeclampsia, we conduct lipidomic profiling of maternal serums of 44 severe preeclamptic and 20 healthy pregnancies from a multi-ethnic cohort in Hawaii. Correlation network analysis shows that oxidized phospholipids (OxPLs) have increased inter-correlations and connections in preeclampsia, while other lipids, including triacylglycerols (TAGs), have reduced network correlations and connections. Thirty-one lipid species from various lipid classes demonstrate predominantly reductions and causal relationships with preeclampsia. They include phosphatidylglycerol (PG), TAG, diacylglycerol (DAG), phosphatidylcholine (PC), cholesterol esters (CE), phosphatidylethanolamine (PE), sphingomyelin (SM), ceramides (Cer-NS), hexosyl ceramides (HexCer-NS), lysophosphatidylcholine (LPC), lysophosphatidylethanolamine (LPE), and free fatty acid (FFA). Many of these lipids are also selected as important features by a linear discriminant analysis (LDA) classifier with high predictive accuracy (F-1 statistic 0.941 and balanced accuracy 0.88), indicating their potential to serve as biomarkers for severe preeclampsia. Our study supports the hypothesis of a phospholipid (PL) centered, dysregulated lipidomic metabolic atlas. That is, severe preeclampsia may be originated from hypoxia, which induces the accumulation of OxPLs through oxidative stress whereas reduces many other lipids (eg. reduced PCs, TAGs and ceramides). These molecular changes coherently lead to dysregulated biological functions, such as insulin signaling and inflammation/infections. Moreover, the lipid changes may also be responsible for the comorbidity between preeclampsia and gestational diabetes, a clinically known risk factor for preeclampsia.

## INTRODUCTION

Preeclampsia is a pregnancy specific syndrome that alJects 2–8% of pregnancies and is diagnosed when a pregnant woman presents with increased blood pressure and proteinuria (Jeyabalan 2013). It is a leading cause of maternal, fetal, and neonatal mortality, especially in low-income and middle-income countries (Saleem et al. 2014). Depending on the onset time, preeclampsia can be categorized as early onset preeclampsia (EOPE, < 34 weeks) and late onset preeclampsia (LOPE). EOPE is a more severe form and is associated with shallow placental implantation into the uterine wall and subsequent placental dysfunction (Raymond and Peterson 2011). Alternatively, preeclampsia can be classified as mild or severe type based on the severity of the symptoms. For severe preeclampsia, the mothers often suffer from potentially fatal pathological manifestations including hypertension, proteinuria, liver rupture, pulmonary edema, and kidney failure (Souza et al. 2013). Moreover, women who had preeclampsia previously have shown 2-3 folds higher risks at developing cardiovascular diseases (CVS) later in life (Leon et al. 2019). The adverse impacts on the fetus include intrauterine growth restriction and preterm delivery. Despite the severity of this condition, few effective treatments are available except expectant management. Delivery of the placenta is the only cure, but this is often coupled with preterm delivery of the fetus (Vigil-De Gracia et al. 2013), with associated life-long health issues in offspring, such as neurodevelopmental disorders and long-term adult onset disorders (Batalle et al. 2012).

Many efforts have been made to systematically understand the biological processes altered in this syndrome, as well as identify potential biomarkers using genomics platforms such as DNA methylation, transcriptomics, proteomics and metabolomics. (Benny et al. 2020; Ching et al. 2015, 2014). However, the systematic changes in lipids, which are more stable compared to other metabolites, are less studied for preeclampsia (S. M. Lee et al. 2019; Olalere, Okusanya, and Oye-Adeniran 2020; Spracklen et al. 2014). Lipids have shown biomarker potentials for many diseases (Balogh et al. 2010; Görke et al. 2010; Han et al. 2007; Pietiläinen et al. 2007; Zhou et al. 2012). Moreover, lipids can reflect the physiological or pathological status of a metabolic disease such as preeclampsia, as they are structural components of cell membranes, signaling mediators, and energy depots. Luckly, lipidomics is an enabling technology to allow for untargeted measurements of hundreds of lipids simultaneously, mostly through liquid chromatography–mass spectrometry (LC-MS) based technology (Han 2016).

Here we conducted untargeted lipidomics profiling of maternal blood from patients with a multi-ethinic cohort of severe preeclampsia (N = 44) patients and those with full-term healthy deliveries (N = 20). By combining LC-MS technology and advanced bioinformatics analysis, we aim to reveal novel insights of lipids and their pathways involved in preeclampsia, as well as providing potential biomarkers for severe preeclampsia. Our results show that a variety of lipids are altered in severe preeclampsia, and some may be direct causes. These molecular changes coherently lead to dysregulated biological functions, such as insulin signaling and inflammation/infections. Oxidized phospholipids (OxPLs) are significantly coordinated and upregulated, presumably due to oxidative stress from hypoxia. Moreover, the commobility between preeclampsia and gestational diabetes, a clinically known risk factor for preeclampsia, may also be mediated through lipids.

## MATERIALS AND METHODS

### Specimens

We obtained samples from RMATRIX Hawaii Biorepository (HiBR), which obtained its own institutional review boards (IRB) approval. 44 maternal plasma samples from clinically diagnosed severe preeclampsia patients and 20 control samples (full-term deliveries) were selected. The clinical summary of the samples is provided in Table 1.

**Table 1:**
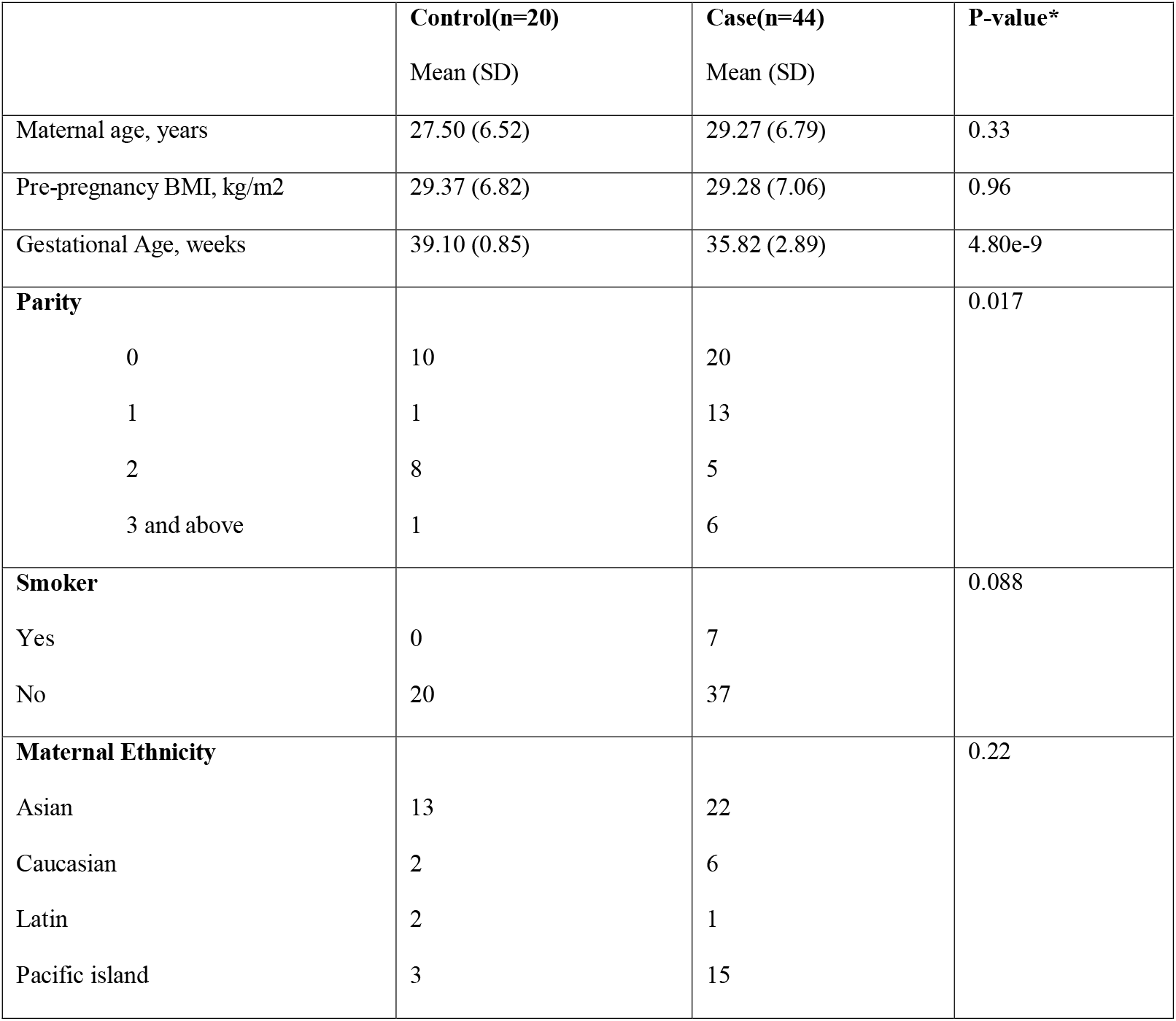
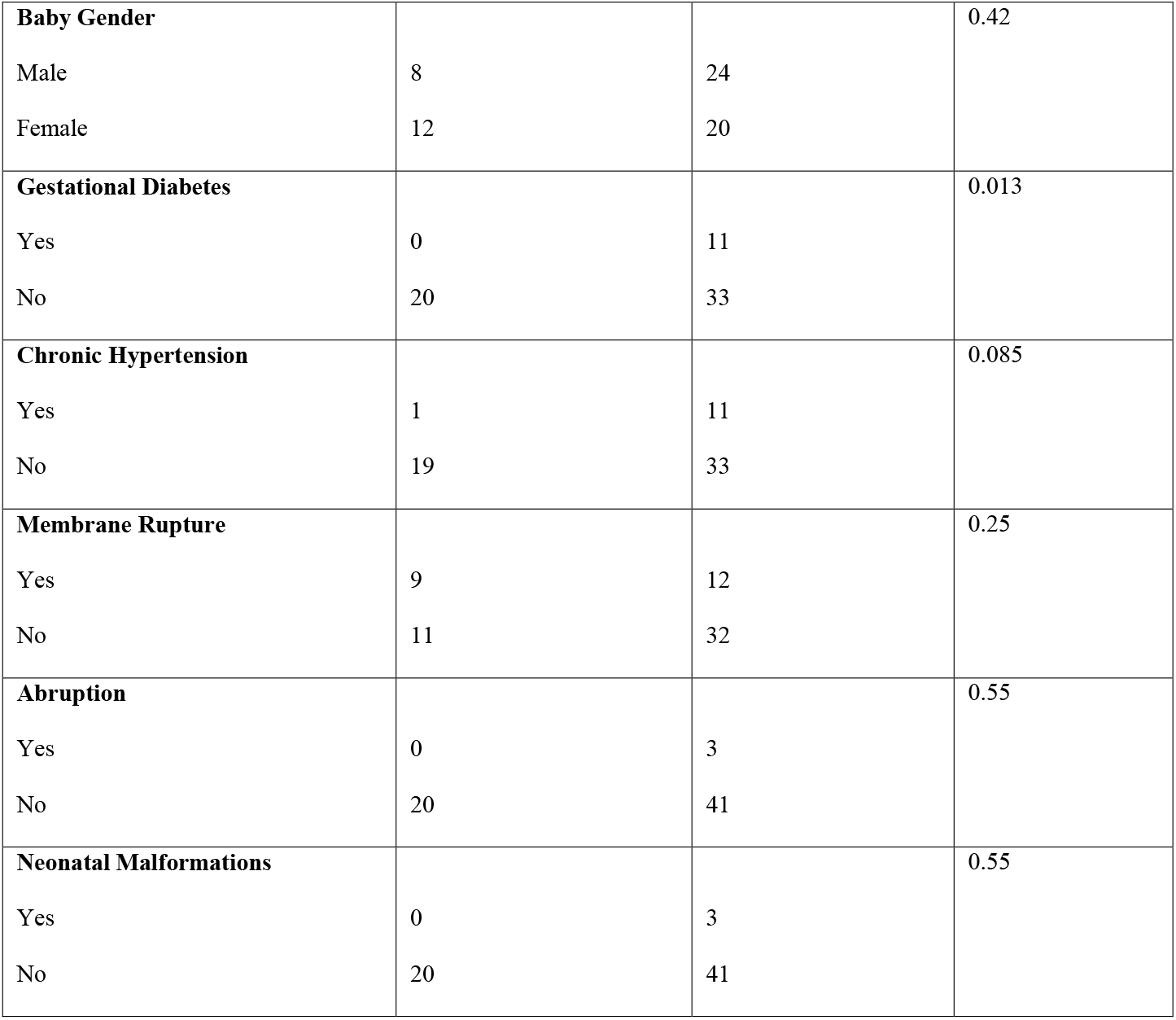
Demographic and clinical characteristics in case and control groups.

### Sample preparation

Plasma samples were stored at − 80 °C until the time of analysis, which was conducted by the Michigan Regional Comprehensive Metabolomics Resource Core. Lipids were extracted using a modified Bligh-Dyer Method (Bligh and Dyer 1959). The extraction was performed using water/methanol/dichloromethane (2:2:2 v/v/v) at room temperature after the addition of internal standards. The organic layer was then collected and dried under a stream of nitrogen before being re-suspended in 100 μL of Buffer B (5%H_2_O:10%ACN:85%IPA containing 10 mM NH_4_OAc) and analyzed using a liquid chromatography tandem mass spectrometry (LC/MS/MS) lipidomics assay (Afshinnia et al. 2016).

### Liquid chromatography/mass spectrometry

The lipid extract was injected onto a 1.8 μm particle 50 × 2.1 mm internal diameter Waters Acquity HSS T3 column (Waters, Milford, MA) that was heated to 55 °C. For chromatographic elution, we used a linear gradient over a 20 min total run time. A 60% Solvent A and 40% Solvent B gradient was used for the first 10 min. Then the gradient was ramped in a linear fashion to 100% Solvent B which was maintained for 7 min. Thereafter, the system was switched back to 60% Solvent B and 40% Solvent A for 3 min. The flow rate used for these experiments was 0.4 mL/min and the injection volume was 5 μL. The column was equilibrated for 3 min before the next injection and ran at a flow rate of 0.400 uL/min for a total run time of 20 min. Data were acquired in positive and negative modes using data-dependent MS/MS with dynamic mass exclusion. Pooled human plasma samples and pooled experimental samples (prepared by combining small aliquots of each experimental sample) were used to control for the quality of sample preparation and analysis (Gika et al. 2008). A randomization scheme was used to distribute pooled samples within the set and a mixture of pure authentic standards was used to monitor instrument performance on a regular basis.

### Lipid identification

Lipids were identified using the LIPIDBLAST computer-generated tandem MS library (Kind et al. 2013). This database contains 212,516 spectra covering 119,200 compounds representing 26 lipid classes, including phospholipids, glycerolipids, bacterial lipoglycan, and plant glycolipids. Quantification of lipids was completed using AB-SCIEX MultiQuant software. Compounds with a relative standard deviation greater than 30% in the pooled samples were excluded.

### Lipidomic data preprocessing

Samples were received in a single batch, with 753 lipid species detected in total. The nomenclature used for individual lipid species begins with the abbreviation of the lipid class followed by the number of carbon atoms in the molecule and then by the number of double bonds. K-nearest neighbors (KNN) method was used to impute missing lipid values, similar to earlier work (Alakwaa, Chaudhary, and Garmire 2018). Data were then log-transformed and subjected to median normalization (Livera et al. 2012), before downstream analysis.

### Source of variation analysis and data screen

The lipidomic dataset of maternal plasma has a total of 753 lipid species. In order to select features capable of distinguishing preeclampsia and control statuses, a preliminary screen was conducted based on the source of variation (SOV) analysis, to explore the contributions of different clinical/physiological factors to the overall lipidomics changes. Only lipid species with a preeclampsia/control F statistic value > 1 were included in further analysis, which meant that for these screened lipids the sample preeclampsia/control status had a regression sum of square (RSS) larger than error sum of square (ESS). This screening process finally identified 292 such lipid species.

### Differential lipid species identification

Wilcox test was used to identify the differentially expressed (DE) lipids between preeclampsia/control status. The lipids with p-values < 0.01 were selected as significant.

### Weighted gene co-expression network analysis

Before weighted gene co-expression network analysis (WGCNA) performance, the normalized lipid values were adjusted via limma (Ritchie et al. 2015). This time, the normalized value of each lipid species was predicted using preeclampsia/control status, and the confounding factors shown in Table 1, including smoking status, baby gender, maternal ethnicity, maternal age, parity, pre-pregnancy BMI, gestational diabetic status, chronic hypertension status, membrane rupture status, abruption, and neonatal malformation status, and then the regression coefficient on preeclampsia/control variable plus the residual value was considered as the adjusted lipid species value, with confounding effects regressed out. Next, the preeclampsia and control samples were separated to different groups, and then analyzed by WGCNA separately (Zhang and Horvath 2005). For both of them, the smallest soft threshold with an R square value > 0.8 was 2, and hence it was chosen to calculate the adjacency score between any 2 lipid species within a sample set. After that, topological overlap value between these 2 lipid species was computed from this adjacency score, as well as their connectivity values. Then, the topological overlap value was further converted to a distance value by subtracting it from 1, providing a distance matrix covering all lipid pairs, which was next used to cluster the lipids using hierarchical clustering and lipid modules could be identified from the resulting dendrogram. Within each module, only lipid pairs with a topological overlap value > 0.5 were kept.

### Classification model

To identify the best machine learning model to distinguish preeclampsia and control samples using lipidomic features, *Lilikoi* was used to construct 7 different machine models (AlAkwaa et al. 2018), including C4.5 decision tree (RPART), gradient boosting (GBM), random forest (RF), elastic net (LOG), linear discriminant analysis (LDA), support vector machine (SVM), and nearest shrunken centroids (PAM). The 64 samples were first split 80/20 into training and test datasets using a 10-fold cross-validation method. The best model is selected based on the F-statistics and blanched accuracy in the testing data set. To determine if adding confounding factors would improve the classifier’s performance, potential clinical confounding factors were also added in addition to the lipid features selected by the best model.

### Lipid pathway and phenotype mapping

To map metabolic pathways to lipid species selected from Wilcox test or classifier, first, their m/z value, ion adduct information, and lipid class information were used as a query to perform a bulk search in LIPID MAPS database. For a query lipid class, all isomers with the same carbon and double bond numbers are returned as the search results. Next, the query lipid and the systematic names of these isomers were used as the input to map to standard HMDB, PubChem, and KEGG IDs in *lilikoi (AlAkwaa et al. 2018)*. These IDs were then used for corresponding pathways analysis.

### Causality analysis

We sorted lipidomics data and clinical features into time series according to the gestational ages of samples. Then we used lmtest package (version 0.9-37) on R platform (version 3.6.3) to perform the Granger causality test for potential causality relationships among lipids and preeclampsia. A causality interaction is significant when p-value < 0.05. Significant causality interactions were collected for further analysis.

### Metabolic enzymes

We identified metabolic enzymes that catalyze the metabolism of lipid species selected from the Wilcox test using Reactome Pathway Database. We analyzed an existing gene-expression data set, namely, GSE44711 in the Gene Expression Omnibus (GEO) database using GEO2R to obtain differential expression of metabolic enzymes in the placenta of patients with preeclampsia (Blair et al. 2013).

### Data availability

The lipidomics data set has been deposited to Metabolomic Workbench, a public repository of metabolomics, under the study ID ST001360. All scripts to analyze the metabolomics data are available at github: https://github.com/lanagarmire/preeclampsa_lipidomics.

## RESULTS

### Overview of the study cohort and lipidomics results

The study is a nested case and control study from a pre-collected population based biobank from University of Hawaii. Maternal plasma of 64 samples (44 severe preeclampsia patients and 20 controls) were used for this study. Table 1 shows the demographic and major clinical characteristics of the subjects. As expected, the patients with severe PE delivered significantly earlier than the controls (averaged gestational age 35.82 weeks vs. 39.10 weeks, p-value = 4.80e-9). Gestational diabetes is also significantly associated with preeclampsia (p-value = 0.013, confirming previous reports that gestational diabetes is a risk factor of preeclampsia (Schneider et al. 2012; J. Lee et al. 2017). In addition, parity is also associated with the preeclampsia group (p-value = 0.017), significantly more samples have a larger parity, as expected. Other risk factors, including smoking and chronic hypertension, show less than significant associations with severe preeclampsia (p-values = 0.088 and 0.085, respectively), which may be due to the limited sample size in this study. Beyond preeclampsia-other clinical factors, we also performed correlation analysis among all clinical factor pairs (Fig. 1A). Supporting results in Table 1, correlations are detected widely between preeclampsia/control status and other variables, such as gestational age (Pearson correlation coefficient, or PCC = −0.533) and gestational diabetes (PCC = 0.307). Gestational diabetes, the other significant clinical factor sharing commodity with preeclampsia, is also correlated with other variables such as BMI.

**Figure 1.**
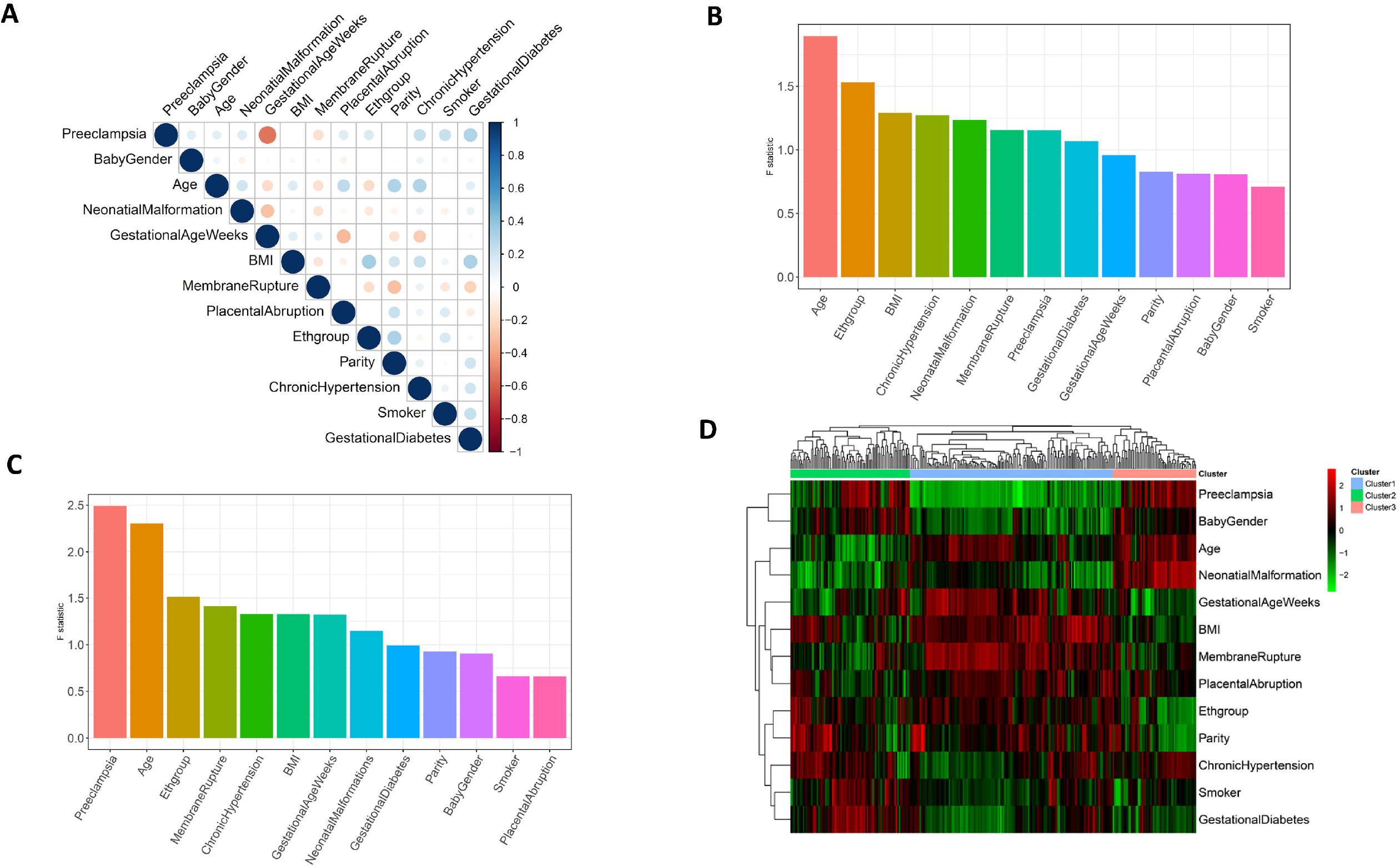
Exploratory analysis of preeclampsia and control samples. (A) Correlation matrix of the 13 phenotypic variables on the 64 samples (20 control vs 44 preeclampsia samples). (B-C) Source of variation (SOV) results across the 64 samples. The error term in the ANOVA model has an F-statistic value of 1. The lipids considered are (B) the 753 lipid species; (C) the 292 lipids, which all have F statistic values of at least 1 for preeclampsia/control category in (B). (D) heatmap showing the correlations between the 292 lipid species and the confounding factors. The rows are lipids, and columns are clinical factors. Each entry of the heatmaps represents the Pearson correlation coefficient value between the lipid and the clinical factor.

The untargeted lipidomic experiments were done in Michigan Regional Comprehensive Metabolomics Resource Core, using a liquid chromatography tandem mass spectrometry (LC/MS/MS) lipidomics assay (Methods). The resulting lipidomics dataset is composed of a total of 753 annotated lipid species. To identify lipids that are truly associated with preeclampsia (rather than due to other confounders), a preliminary screen was conducted on the lipid species using a source of variation (SOV) analysis. 292 lipid species with preeclampsia/control F statistic values > 1 were selected for subsequent analyses. As a confirmation, the ranking of preeclampsia/control status is improved from the 7th in the whole dataset (Fig. 1B) to the 1st in the filtered metabolomics subset with 292 lipids (Fig. 1C).

We next examined the correlations between the clinical variables and the 292 screened lipid species by heatmap (Fig. 1D). Hierarchical clustering analysis on the 292 lipid species shows 3 main clusters. Cluster 1, 2 and 3 are composed of 146, 86 and 60 lipid species, respectively. Cluster 2 is significantly enriched in diacylglycerol (DAG) lipids (Fisher’s p-value = 3.66e-2, odds ratio = 2.20). Cluster 3 has a large enrichment in 2 kinds of oxidized phospholipids (OxPLs): oxidized phosphatidylethanolamine (OxPE, Fisher’s p-value = 2.55e-5, odds ratio = 4.87) and oxidized phosphatidylcholine (OxPC, Fisher’s p-value = 1.56e-3, odds ratio = 4.38), but deprivation of triacylglycerol (TAG, Fisher’s p-value = 6.32e-3, odds ratio = 0.125). No lipid species is detected as significantly enriched in Cluster 1. In summary, the results provide the initial evidence that lipidomics are associated with severe preeclampsia, despite the complexity due to other clinical variables. The complex correlations between preeclampsia and other clinical variables suggest that their relationships may be mediated through molecules including lipids studied here.

### Correlation network analysis of lipidomics in relation to preeclampsia

To elucidate the relationships between lipidomics and severe preeclampsia, we next constructed the correlation networks for preeclampsia and control samples, using weighted gene correlation network analysis (WGCNA) method on the 292 screened lipid species (Zhang and Horvath 2005). In both preeclampsia and control conditions, the network contains 2 modules, a large one and a small one (Fig. 2A and 2B). Each of them corresponds to the counterpart well between preeclampsia and control group, as the common lipid species have significant overlap between the 2 conditions (Fig. 2C). The small network module is more densely connected in PE (0.64) than the controls (0.56), with enrichment of OxPE and OxPC, two types of OxPLs, in both conditions. However, the enrichments of OxPE and OxPC in preeclampsia (Fisher’s p-values are 8.81e-9 and 1.70e-4, respectively) are also much more significant in preeclampsia condition, compared to the controls (1.49e-6 and 1.40e-2, respectively, in control), as shown in Fig. 2D. OxPLs have been previously associated with a variety of diseases, including arteriosclerosis, diabetes, and cancer (Aoyagi et al. 2017). The increase in both the enrichment of OxPLs as well as their interconnections suggest that preeclampsia is another OxPLs related disorder. On the other hand, the large network module has less connection density in preeclampsia (0.26) than in controls (0.36), with evidence of prevalence of TAG and DAG. In fact, TAG (Fisher’s p-value = 1.73e-2) and DAG lipids (Fisher’s p-value = 1.95e-2) are both significantly enriched in preeclampsia, whereas in controls only TAG enrichment is significant (Fisher’s p-value = 3.32e-2), but not DAG (Fisher’s p-value of 8.87e-2).

**Figure 2.**
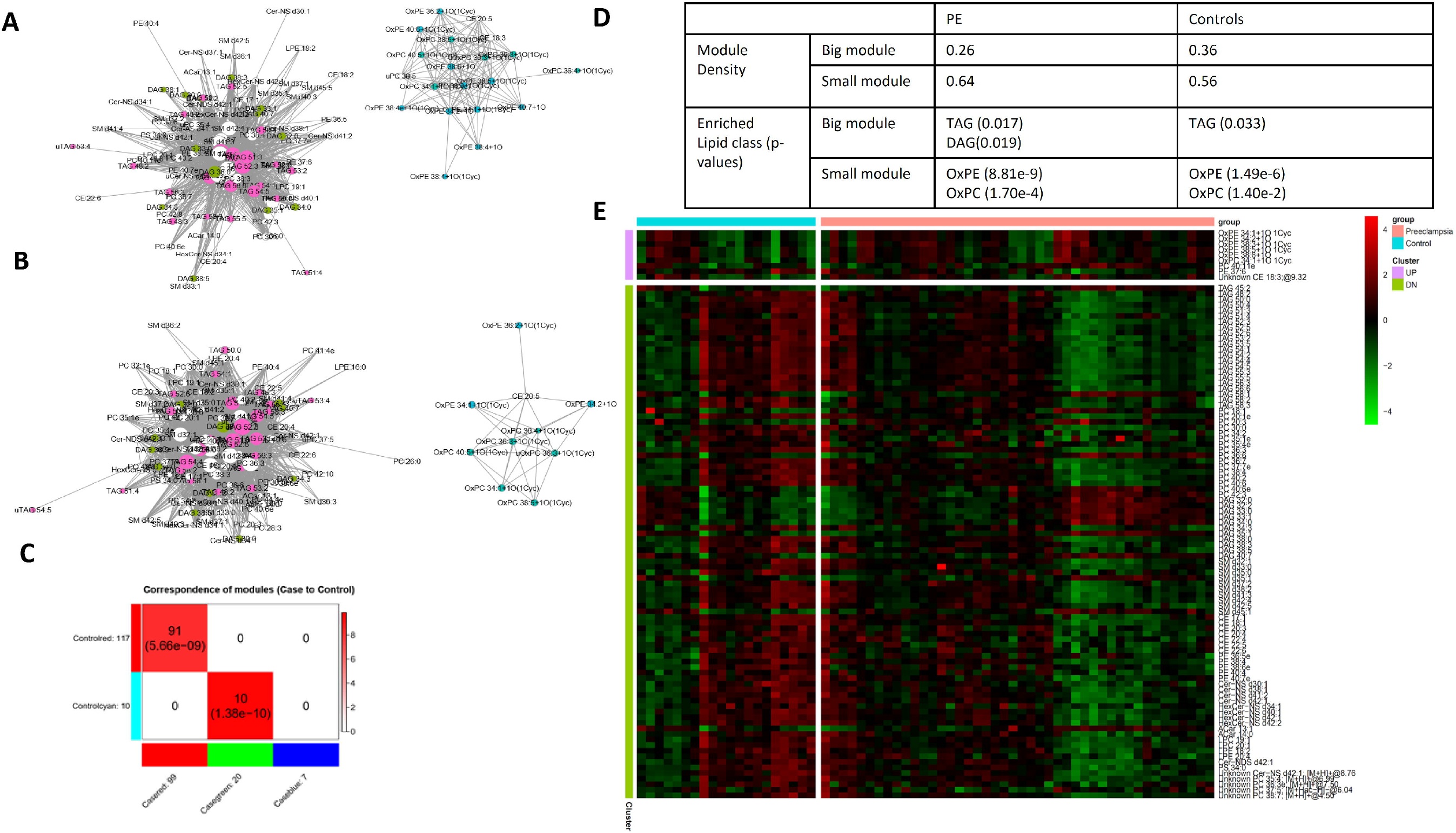
WGCNA network comparison between preeclampsia and control samples. (A-B) WGCNA network in preeclampsia (A) and controls (B), respectively. Each node represents a lipid species, whose size is proportional to the node connectivity value in a WGCNA network. (C) Overlap between modules of networks in control and in preeclampsia samples. (D) Table showing more properties of modules in networks of control vs preeclampsia samples, including module density and enriched lipids in each module. (E) Heatmap of lipids with significant connectivity difference in WGCNA networks (A-B), between control vs preeclampsia. Connectivity value is defined as increased or decreased, if (the connectivity value in the preeclampsia network - connectivity value in the control network) is larger than 5 or less than −5, respectively.

To further explore the lipid correlation differences between preeclampsia and controls, we specifically extracted lipid species with significant changes in network connectivity values, meaning that the difference of the sums of all edge weights from a node between case and control conditions are greater than 5 (upregulated lipids) or less than −5 (downregulated lipids). With such stringent cutoff threshold, 9 lipids are identified upregulated and 92 are downregulated (Fig. 2E), and TAG and DAG are the only 2 enriched lipid classes in the downregulated group (Fisher’s p-value = 1.42e-2 and 4.53e-2). The decrease in both the levels (Fig 2E) and the coordination (Fig 2C) of these lipids suggest that their biogenesis processes are disrupted in preeclampsia. Of the 9 lipids with increased intensities, six are OxPE or OxPC lipids, corroborating the earlier WGCNA results on increased correlations and connections among them (Fig 2A-C). Together, these results show that oxidative lipid genesis is enhanced, whereas TAG and DAG lipid metabolism is dys-regulated in severe preeclampsia.

### Lipids and their pathways associated with preeclampsia

In addition to correlation network analysis, we also analyzed the overall lipid concentration changes between the two conditions using Wilcox rank tests. 31 lipid species are identified as significantly different with p-values less than 0.01. Four lipids are upregulated: free fatty acid (FFA) 20:1, phosphatidylcholine (PC) 36:6e, phosphatidylethanolamine (PE) 37:2 and phosphatidylglycerol (PG) 34:1. Twenty-seven lipid species are downregulated (Fig. 3A, Supplementary Table 1). Among the 27 downregulated lipids, 17 also have reduced network connectivities in WGCNA, suggesting their dual attenuations on both lipid concentrations and correlations. To identify the subset of differential lipids among the 31 lipids that are only due to preeclampsia but not any other clinical conditions (eg. gestational diabetes), we conducted confounder stratification and identified 8 lipids. They are lysophosphatidylethanolamine (LPE) 18:2, lysophosphatidylcholine (LPC) 20:5, LPC 18:2, PC 18:2e, sphingomyelin (SM) d36:1, ceramide (Cer-NS) d30:1, LPE 20:4, and PE 37:2.

**Figure 3.**
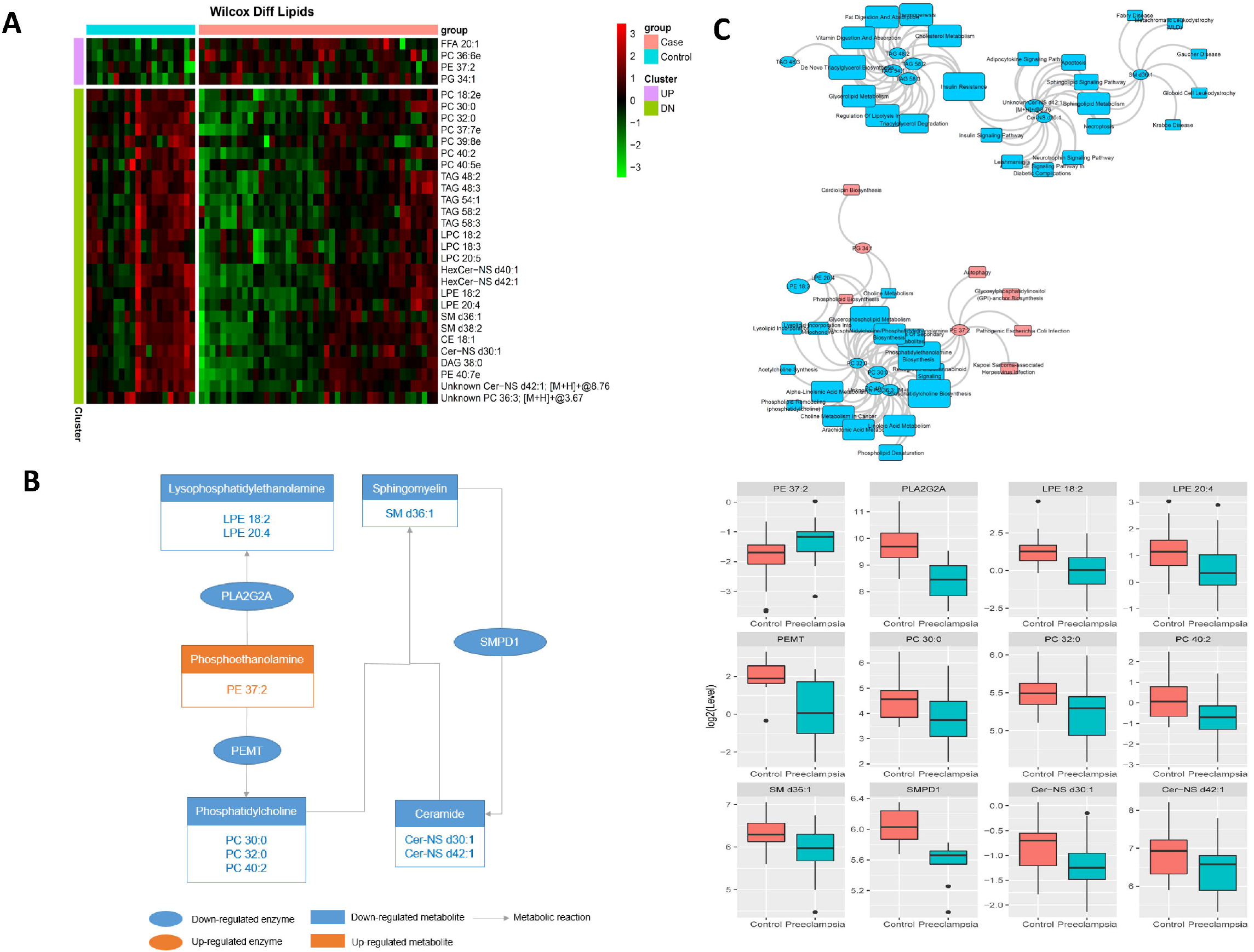
Lipids show significantly different levels in preeclampsia and control samples. (A) Heatmap of lipids with significant difference between preeclampsia and control samples. (B) A module of coherent lipid changes explained by their metabolomic reactions. The color and shape annotations are shown in the figure. The corresponding levels of respective enzymes and metabolites between preeclampsia and control samples are shown in the box plots on the right. (C) Bipartite graph of lipids in (A) and their affiliated metabolic pathways. Elliptic nodes: lipid. Rectangular nodes: pathways from HMDB, PubChem and KEGG database. Blue color: downregulation in preeclampsia. Pink color: upregulation in preeclampsia. Note: lipids without any metabolomic pathway affiliations are omitted.

Some of these significant lipid classes are linked tightly during their synthesis, which may explain the coherent changes. For example, PE 37:2 (upregulated lipid) is the synthetic precursor for 6 of the downregulated lipids (LPE 18:2, LPE 20:4, PC 30:0, PC 32:0, PC 36:3, and PC 40:2). Enzyme PLA2G2A catalyzes PE 37:2 to the LPEs, whereas PEMT converts it to PCs (Fig. 3B). Supporting our lipidomics results, gene expression results in another preeclampsia and control placenta data (Blair et al. 2013) also show these two enzymes are downregulated in preeclampsia (PLA2G2A, log2FC = −1.41, p-value = 5.47e-3; PEMT, logFC = −1.75, p-value = 5.81e-2), agreeing with the accumulation of PE 37:2 and decreased LPEs and PCs levels in our data. As PCs are synthetic precursors of SM d36:1, which can be further converted to ceramides (eg. Cer d30:1 and Cer d42:1), it also explains the reduced levels of another three lipids SM d36:1, Cer d30:1 and Cer d42:1 (Fig. 3B).

To understand the functions of the lipids with significantly different concentrations in preeclampsia vs. control, we attempted pathway enrichment analysis. However, this task was not easy, as the current shotgun lipidomic technique can only identify lipids by their class group and total number of carbons and double bonds, rather than providing definitive and unique identifications. To overcome this issue, we performed isomer searches of each lipid class first, then used the lipid and isomers together to search for corresponding HMDB, PubChem, and KEGG pathways. As a result, 16 out of 31 lipid classes yielded corresponding pathways (Fig. 3C). A group of ceramides are overall reduced in preeclampsia samples. Ceramides are part of various signaling pathways, including sphingolipid pathway, adipocytokine signaling pathway and insulin signaling pathway, which are involved in diabetes. Therefore, the comorbidity of preeclampsia and gestational diabetes may be linked through ceramide and TAG lipid class (Fig. 3C). PCs (eg. PC 30:0, and PC 40:2) are overall reduced in preeclampsia, and they are linked to suppressed arachidonic acid metabolism and linoleic acid metabolism pathway. This is consistent with previous observation of arachidonic acid production disturbance in preeclampsia (Walsh and Parisi 1986), and supports the claim of potential prevention effect on this disease by linoleic acid supplementation (M and Herrera M 1998). On the other hand, as one of the few lipids increased in preeclampsia, PE 37:2 is linked to various infection and immune response pathways, including “pathogenic E. Coli infection” and autophagy process.

### Metabolomics based preeclampsia biomarker model

An important application of lipidomics data is to screen for diagnostic biomarkers for diseases. To do so, we split samples with 80/20 ratio to training and testing data. After feature selection using a ciretera of information gain value greater than 0.1, we compared the performance of several popular machine learning algorithms in *Lilikoi* R package in the testing data, to obtain the best classification method for the lipidomics data (Fig. 4A). These classification algorithms are: linear discriminant analysis (LDA), random forest (RF), elastic net (LOG), gradient boosting (GBM), support vector machine (SVM), nearest shrunken centroids (PAM), and classification tree (RPART). We used F1 statistics and blanched accuracy metrics to evaluate the models, given the unbalanced size of the preeclampsia and controls. We chose LDA as the final model, as it yields the largest F1 statistic of 0.941 in the testing dataset, as well as the highest balanced accuracy 0.88 (Fig. 4A).

**Figure 4.**
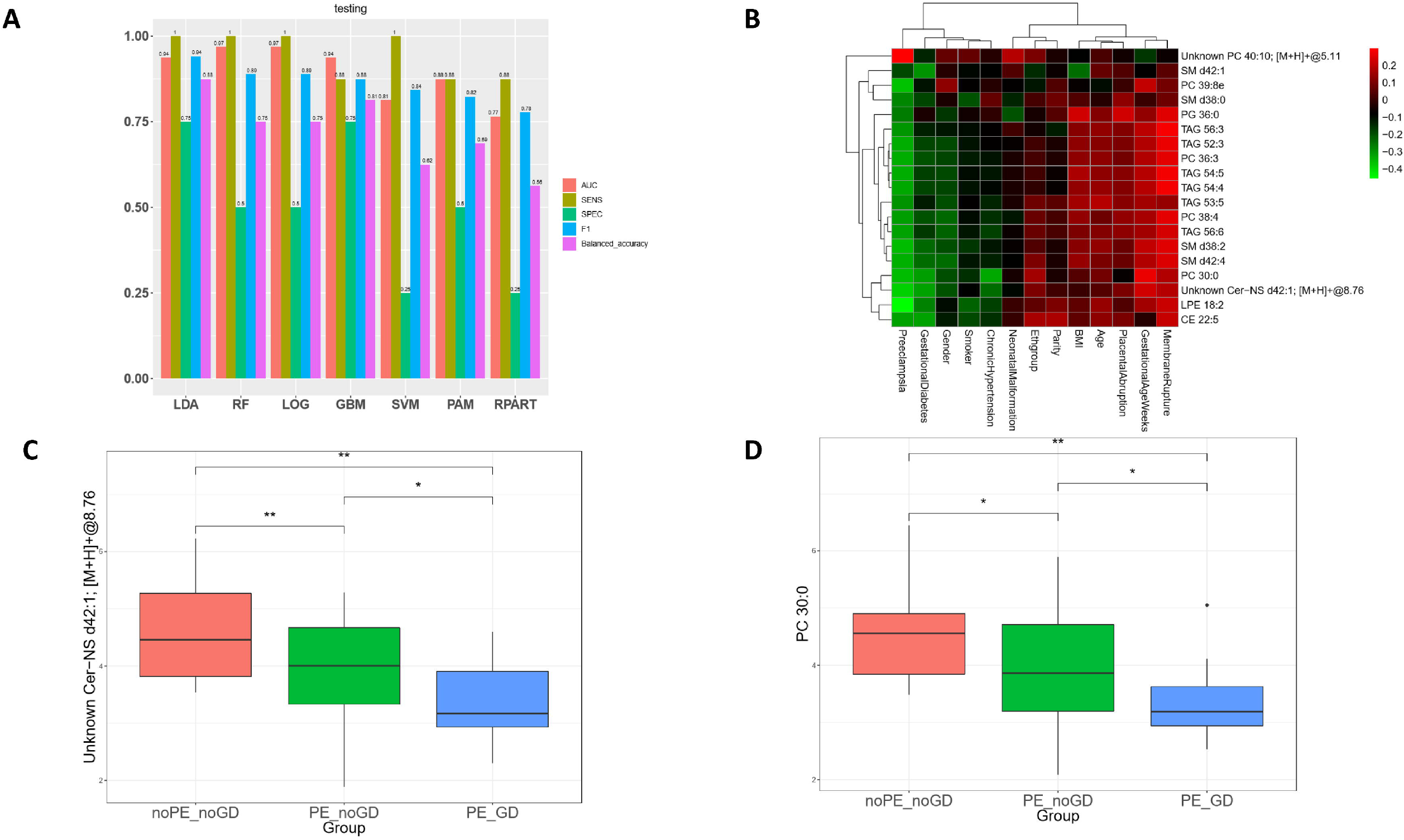
Biomarker classification model for preeclampsia. (A) Comparison of seven popular classification models using metabolomics data on the hold-out testing data, from left to right: LDA, RF, LOG, GBM, SVM, PAM and RPART. (B) Heatmap of correlation coefficients between 19 lipid features of LDA and other clinical variables. (C-D) Selected lipid features from the LDA classifier, showing significant differences in control, preeclampsia and gestational diabetes. (C) Cer-NS d42:1 and (D) PC 30:0 *: p-value < 0.05, **: p-value < 0.01, t-test.

Given the potential confounding from some clinical variables (Fig. 1), we also checked if adding other clinical variables as additional features can improve the prediction performance of LDA, but the AUC of precision-recall curves does not change (Supplementary Fig. 1), indicating that the additional clinical variables do not independently contribute to the model prediction aside from metabolomics features. For the LDA model, it includes 19 lipid features (Fig. 4B), and all of them have negative correlations with preeclampsia except PC 40:10. Putting back into the context of the 292 lipids, 17 of these lipid biomarkers belong to Cluster 1 in Fig. 1D, where the lipids are predominantly negatively correlated with preeclampsia, while the other 2 lipids (PC 40:10 and SM d42:1) belong to Cluster 3 in Fig 1D. Additionally, some lipid biomarkers are significantly associated with both preeclampsia and gestational diabetes (Fig. 4C-D). Cer-NS d42:1 levels are significantly lower in preeclampsia patients without gestational diabetes complication (p < 0.01), compared to those in controls (Fig. 4C); it is furtherly reduced among preeclampsia patients with gestational diabetes (p < 0.05). Another lipid, PC 30:0 is also a biomarker feature with very similar trends of changes with both conditions (Fig. 4D).

### Predicted causality interactions among lipids and preeclampsia

To explore the potential causal relationship between lipids and preeclampsia, we performed Granger causality test (Geweke 1982) and predicted significant causality interactions (p-value < 0.05). The results show the causality interaction from ceramide Cer-EOS d49:1, Cer-EODS d55:2, Cer-NS d43:2, Cer-NS d40:1, Cer-NDS d36:0 and Cer-NDS d42:2, phosphatidylethanolamine PE 36:5e and PE 36:1e, phosphatidylcholine PC 44:4, PC 35:1e, PC 36:3e and PC 36:3e, oxidized phosphatidylcholine OxPC 36:4+1O(1Cyc), sphingomyelin SM d40:4, SM d34:0, SM d40:5 and SM d38:3, cholesterol ester CE 20:3 and CE 18:2, triacylglycerol TAG 44:0, TAG 52:4 and free fatty acids FFA 22:0 to preeclampsia, respectively (Fig. 5). The causality test also shows the causality interaction from preeclampsia to phosphatidylcholine PC 39:8e and phosphatidylethanolamine PE 33:2. It’s interesting to see causality interactions from phosphatidylethanolamines of larger molecular weights (PE 36:5e and PE 36:1e) to preeclampsia, and then from preeclampsia to phosphatidylethanolamine of smaller molecular weight (PE 33:2).

**Figure 5.**
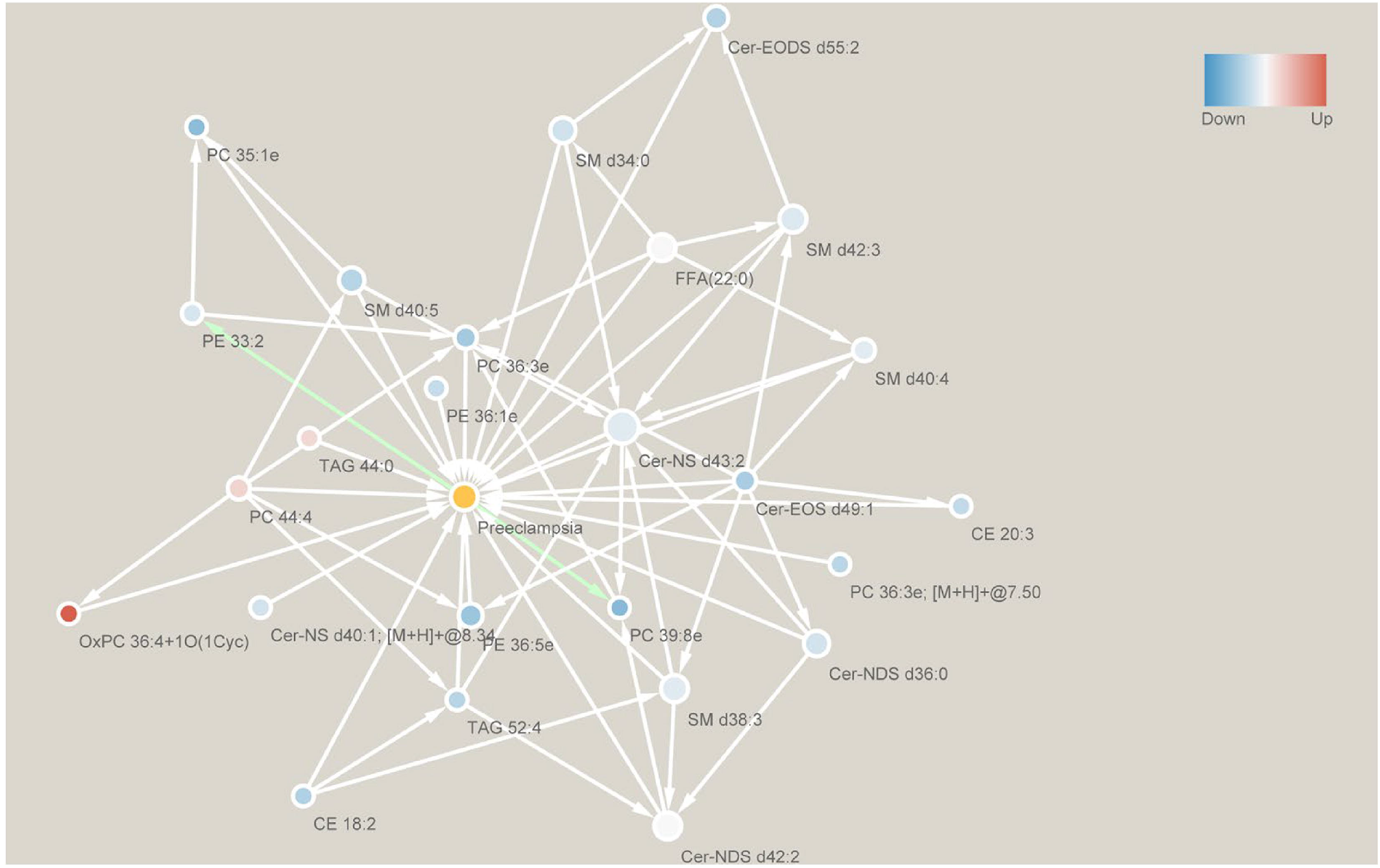
Predicted causality interactions among lipids and preeclampsia. Edge goes from cause to result. White edges go towards preeclampsia, while green edges start from preeclampsia. Blue nodes are down-regulated lipids, while red ones are up-regulated lipids. Only significant (p-value < 0.05) causality interactions are shown.

## DISCUSSION

Preeclampsia is a complex, heterogeneous disorder of pregnancy (Jeyabalan 2013). To improve our understanding of severe preeclampsia, we conducted a lipidomics study on a unique multi-ethnic cohort in Hawaii.

One of the most significant and novel findings in this study is the increased coordinations and intensities among OxPE and OxPC lipids in preeclampsia. Similar increases of OxPLs have been observed and associated with various other diseases, such as arteriosclerosis, diabetes, and cancer (Aoyagi et al. 2017). OxPLs have a known close relationship with oxidative stress. In preeclampsia, hypoxia is one of the most important features and a source of oxidative stress (Freikman et al. 2008). This hypoxic condition in preeclampsia is derived from the incomplete remodeling of spiral arteries by extracellular trophoblasts in this disease. Furthermore, such spiral arteries cannot provide adequate blood to the placenta, resulting in a hypoxic condition (Pijnenborg, Vercruysse, and Hanssens 2006). Thus we speculate that enhancement of OxPLs coordination is the result of oxidative stress induced by hypoxia.

Moreover, the OxPL change may further lead to other lipidomic profile changes observed, such as the reduced TAG lipids and their coordinations. This is due to the fact that phospholipids (PLs), the precursors of these OxPLs, are also sources for converting DAG to TAG. PLs have a central role in lipid metabolism. In healthy individuals, DAG and PL can be used by the cell to produce either TAG, or lysophospholipids and fatty acids, including arachidonic acid (via lecithins and linolenic acid metabolism). Under the hypoxic condition of preeclampsia, many PLs are oxidized to OxPLs, reducing their availability to be used for other lipids downstream, such as TAG or ceramides, which we observed the reduction in preeclampsia in Fig 3A. The data suggest the lipidomic model of preeclampsia, where hypoxia may lead to increased OxPL levels due to oxidative stress, and further the reduction of TAG etc. (Fig. 6).

**Figure 6.**
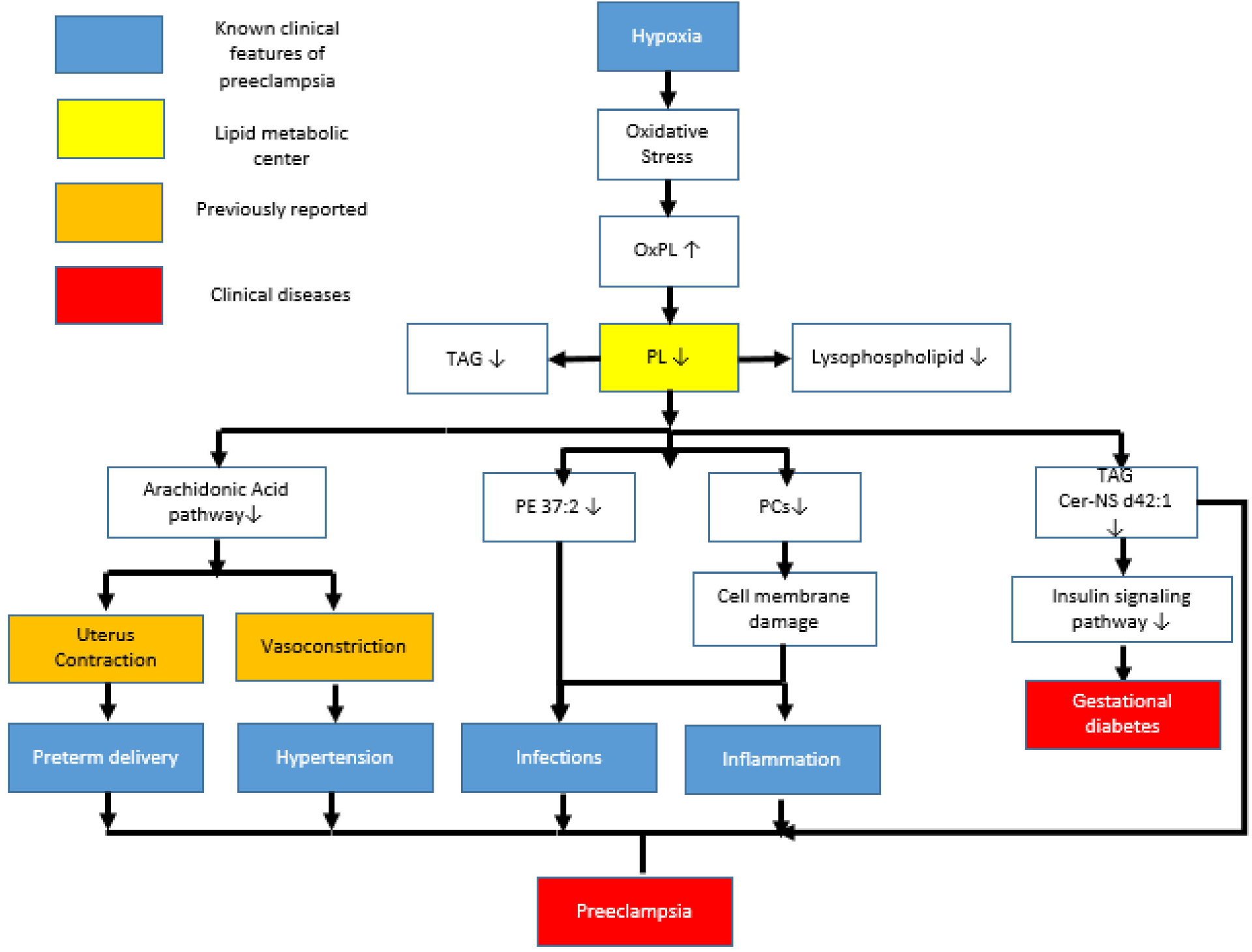
A proposed physical model of lipidomic changes in severe preeclampsia. A hypoxia induced, PL centric model of dysregulated lipid metabolism in preeclampsia.

Changes in several lipid classes may contribute to inflammatory stress observed in preeclampsia. The first link is from the increase of PE 37:2, which mediates the pathways of pathogenic E. Coli infection as well as autophagy process (Fig 3C), both of which have a close relationship to inflammation (Tanida, Ueno, and Kominami 2008). This lipid can specifically interact with the bundle-forming pilus (BFP) of pathogenic E. Coli for bacterial autoaggregation and adherence to host cells, and contribute to pathogenic E. Coli. infection (Khursigara et al. 2001). PE 37:2 is also involved in autophagy. During this process, the microtubule-associated protein 1A/1B-light chain 3 (LC3) in cytosol can conjugate to it to form LC3-phosphatidylethanolamine conjugate (LC3-II), which is recruited to autophagosomal membranes (Tanida, Ueno, and Kominami 2008). LC3-II was shown to increase in placenta, indicating an increased autophagic activity during the pathogenesis of this disorder (Oh et al. 2008). Secondly, many PCs are shown significantly reduced in severe preeclampsia (Fig. 3A). PCs are the main phospholipids of cell membranes (up to 50%), and their downregulation likely causes cell membrane damage, which may contribute to maternal/fetal tissue injuries and increase the risks of infections and inflammation. Moreover, PCs are linked to down regulation of arachidonic acid and linoleic acid metabolism, both of which are related to inflammation. In particular, arachidonic acid metabolism is of great importance in preeclampsia due to its central role as the inflammatory mediator (Walsh and Parisi 1986). Arachidonic acid is a precursor of prostaglandins, which are largely pro-inflammatory. Cyclooxygenase 1/2 (COX1/2) converts arachidonic acid to prostaglandin H2 (PGH2), which can further be converted to other prostaglandins. Aspirin, which is recommended to use at a low-dose to prevent or delay the onset of preeclampsia by American College of Obstetricians and Gynecologists, blocks the activity of COX1/2 irreversibly.

Lipid dysregulation may also explain the commobility between preeclampsia and gestational diabetes, with several lines of evidence. For example, the pathway analysis has pointed to changes of TAGs and ceramides, hinting to disturbed insulin signaling pathway activity (Fig. 3C), which are well known responsible for diabetes. In particular, Cer-NS d42:1 is an important ceramide with definite associations to insulin signaling (Fig. 3B). It is also selected as a biomarker for preeclampsia (Fig 4B) due to its significant changes (Fig. 3A). Another lipid, PC 30:0 is also a biomarker feature significantly associated with preeclampsia (Fig. 3A). It also significantly reduced further within preeclampsia patients who have gestational diabetes. In fact, among the 19 biomarkers selected by the classification model for preeclampsia, most of them have very similar correlations between preeclampsia and gestational diabetes. Thus, the commodity between preeclampsia and gestational diabetes are tightly coupled from the lipid metabolism aspect.

Comparisons among our study with previous lipidomics preeclampsia studies reveal certain degrees of consistency. In an earlier study on plasma of 10 pregnant women with preeclampsia and 10 controls, TAG and other neural lipids such as sterol showed a lower level in the disease condition (Korkes et al. 2014), consistent with our results. Another lipidomics biomarker study was conducted on serums of mostly caucasians collected in 12–14 weeks pregnancies, from a discovery set of 27 controls and 29 preeclampsia patients, and a validation set of 43 controls and 37 preeclampsia cases (Anand et al. 2016). They identified 23 potential biomarkers, among them the largest lipid class was PCs, similar to us. They also identified TAGs, CEs as well as oxidized lipids (eg. OxPC, OxSM, OxCE) similarly. Some molecules consistently showed up in the biomarker panels, with m/z values of 283 (fatty alcohol and aldehydes, oxidized cholesterol, unknown identity, unknown putative constituents), 383 (oxidized cholesterol, unknown identity, unknown putative constituents), 445 (Cholesterol derivatives, unknown identity, unknown putative constituents), 645 (CE 18:4) and 784 (PC 36:3). One major difference between the two assays is that our study has more annotated lipids, which helps to reconstruct the lipidomic landscape and understand the metabolic mechanisms better. Another study focusing on circulating sphingolipids in the plasma drawn at the first, second, and third pregnant trimesters of 7 preeclampsia and 7 control samples, showed that some sphingomyelin and ceramide lipids were significantly down-regulated in the preeclampsia group (Dobierzewska et al. 2017), consistent with our observations. Interestingly, a study on placental lipid profiles from 23 preeclampsia pregnancies showed higher neutral lipid contents than 68 healthy controls (40% higher triacylglycerol and 33% higher cholesteryl ester), as well as increases in most PC lipid species. The authors concluded that placenta has a lipid storage status under preeclampsia condition (Brown et al. 2016). Their result in placentas is almost completely complementary to our maternal blood observation, suggesting a “source and sink” scenario at play (Xie et al. 2015). That is, the deprivation of these lipids in the blood supplies the lipid storage in preeclamptic placentas.

Our study highlights the lipidomic changes manifested in severe preeclampsia and points to plausible lipid metabolic mechanisms. We have identified a close relationship between OxPLs and preeclampsia, presumably through the oxidative stress due to hypoxia. We propose that many significant blood lipid changes (eg. reduced TAG, PCs, and ceramides) observed in the severe preeclampsia patients can be explained by the PL centered lipidomic axis. These molecular changes coherently mediate dysregulations in biological functions, such as insulin signaling and inflammation/infections. Moreover, these changes also partially explain the commobility between preeclampsia and gestational diabetes, a clinically known risk factor for preeclampsia. The selected lipid features for preeclampsia may have the potential to serve as diagnostic biomarkers and will be of interest for post COVID follow-up validation studies.

## Data Availability

The lipidomics data set has been deposited to Metabolomic Workbench, a public repository of metabolomics, under the study ID ST001360.

## ACKNOWLEDGEMENT

LXG is supported by grants K01ES025434 awarded by NIEHS through funds provided by the trans-NIH Big Data to Knowledge (BD2K) initiative (www.bd2k.nih.gov), R01 LM012373 and LM012907 awarded by NLM, R01 HD084633 awarded by NICHD.

## Supplementary Figures and Tables

Supplementary Fig 1. The Precision-Recall ROC curves of lipid only model, clinical confounder only model, and the combined model of lipids and clinical confounders.

